# Effectiveness and Cost-effectiveness of Home-Based Occupational Therapy with Activity Monitoring for Daily Functioning of People Post-Stroke: Protocol for a Partially Randomised Usual Care Stepped-Wedge Trial

**DOI:** 10.1101/2025.10.12.25337833

**Authors:** Sanne Pellegrom, Margriet C. Pol, Ton Satink, Margo van Hartingsveldt, Johanna Maria van Dongen, Gerben ter Riet, Bianca M. Buurman, Maud Graff

**Author notes:** Data availability: All relevant data from this study will be made available upon request after study completion.

## Abstract

**Background:** Transitioning from a geriatric rehabilitation centre or hospital to home is challenging for persons post-stroke, as they must increase their ability to perform daily activities and self-manage their lives. Moreover, persons post-stroke exhibit more sedentary behaviour at home than during rehabilitation, which is detrimental to their health. The Occupational Therapy at Home E-Rehabilitation (OTHER) intervention aims to increase daily activities and self-management in persons post-stroke. OTHER involves in-person (online) coaching with information from activity monitoring. Here, we describe the design of a study investigating the (cost-)effectiveness of OTHER on the recovery of daily activities among older persons after stroke and evaluate its implementation process.

**Methods:** A partially cluster randomised stepped-wedge trial is conducted in nine geriatric rehabilitation centres and two hospitals, aiming to include 171 post-stroke participants (aged ≥ 60 years) who are starting (inpatient) rehabilitation to return home. The intervention group receives the OTHER intervention by an occupational therapist, starting at the institute and continuing at home. This intervention consists of activity monitoring to increase insight into movement behaviour, as well as coaching by occupational therapists on improving daily activities at home. This supports persons post-stroke in performing daily activities and increasing self-management. The control group receives care as usual. The primary outcome is the performance score of the Canadian Occupational Performance Measure, which measures a person’s perceived daily functioning at 6 months. A process evaluation will be conducted to assess the implementation process of the OTHER intervention during the research period.

**Discussion:** This trial is studying the effect of activity monitoring combined with home-based occupational therapy coaching for persons post-stroke. With this study, we aim to provide insight into the effectiveness and cost-effectiveness of the OTHER intervention compared to care as usual for persons post-stroke, and describe the process of implementation, which includes home-based geriatric rehabilitation.

**Trial registration:** ClinicalTrials.gov: NCT05855226, and Organization’s Unique Protocol ID: 80-86900-98-022, registered may 11^th^ 2023

## Introduction

Stroke is a leading cause of disability and the third-largest contributor to disease burden globally. In the Netherlands, around 40,000 individuals experience a stroke annually, accounting for 2.5% of healthcare costs (1–4). Outcomes vary widely, from minor impairments to long-term disabilities, depending on stroke location and severity (2,4). Persons post-stroke often face a prolonged, personal process of adaptation to limitations in motor, cognitive, and emotional functioning (5). Stroke may also affect partners and family members, who often take on the role of informal caregiver and may experience emotional and physical burdens themselves (6,7).

After hospital discharge, many older stroke survivors require geriatric rehabilitation (GR), often delivered in specialised rehabilitation centres, to regain functional abilities and adapt to daily life (8–10). The rehabilitation goals are often focused on regaining independence (9). However, persons post-stroke are frequently restricted in their daily functioning and depend on support from both professionals and informal caregivers during rehabilitation in transitioning to go home. Continuation of GR at home is therefore crucial for resuming daily life and daily functioning (11). Despite intensive GR during the transition from geriatric rehabilitation or hospital to home, people are often unprepared to self-manage (5).

Physical inactivity and sedentary behaviour are common post-stroke, including light activities such as walking or household tasks, which are often not performed (12,13). Yet, performing light physical activity is linked to better health outcomes (12). Performance of daily activities is essential to restore functioning and develop self-management skills. Effective self-management programs are often home-based and involve informal caregivers (14). Support from caregivers, professionals, and activity monitors that offer real-time feedback may help increase motivation and improve activity engagement (11) and, as a result, enhance self-management skills. Given the rising demand for healthcare services due to population ageing and a shrinking healthcare workforce, e-rehabilitation interventions could be a cost-effective solution to support home-based GR for persons post-stroke (15). Studies suggest that e-health interventions are as effective as face-to-face interventions (16) and can support self-management, autonomy, improve daily activity performance, and facilitate continuity of care (17,18). Additionally, commercially available mobile and web applications, as well as activity sensors, are being increasingly utilised in stroke rehabilitation, which helps to raise awareness and provide feedback to patients that encourages daily activity (19). Promoting daily activity may also help counteract inactivity and sedentary behaviour (5,15).

However, despite the growing availability and promising effects of these technologies for functional improvement after stroke (20,21), there is limited evidence on their use in home-based rehabilitation for increasing daily activities for persons post-stroke.

In particular, the Home E-Rehabilitation (OTHER) intervention—an intervention combining activity monitoring with hybrid coaching delivered by an occupational therapist— was inspired by an intervention that has been shown to improve daily functioning in people after hip fracture (22). These intervention components seemed feasible for use in a person-centred occupational therapy intervention for people post-stroke, as described earlier in a paper of the feasibility study of OTHER (under submission). However, their effectiveness and cost-effectiveness have not yet been evaluated in persons post-stroke.

This paper describes the study protocol for an effectiveness and cost-effectiveness trial with a process evaluation, evaluating the impact of OTHER on improving daily functioning and self-management in activities of daily living and quality of life among community-dwelling persons post-stroke, compared to care as usual, at six-month follow-up, and evaluates its implementation process.

## Materials and Methods

Before outlining the trial design in detail, we wish to be transparent about the current status of the trial. Recruitment began on March 23, 2023. However, by April 2024, enrolment was much lower than expected (N = 57 April 2024; we expected over 100 participants). In response, we implemented two key adjustments: 1) two GR centres and two hospitals were added, and 2) recruitment was extended by nine months and will end at the end of July 2025. The following sections outline the original plan and the adjustments made, the Consolidated Standards of Reporting Trials guidelines for stepped wedge cluster randomised trials (23) and the SPIRIT checklist were used (see S2).

### Trial design

The study is a partially cluster randomised stepped-wedge trial. The intervention was developed and tested following the steps of the first two phases of the MRC framework (Pellegrom et al, Manuscript submitted for publication). This study contains the third phase of the MRC framework, following the steps of the evaluation phase (24,25)).

Table 1 provides an overview of the trial design. Initially, the clusters contained the nine GR centres. Every cluster begins with care as usual (see Table 1), during which the data will be collected for the control group. Every three months, a cluster will initiate the OTHER intervention, and all participating occupational therapists in the cluster will perform the OTHER intervention with new participants after stroke with the OTHER intervention (see Table 1). Data will be collected during the intervention period. Each cluster continues the OTHER intervention till the end of the trial. However, two hospital settings and two GR centres were also added after one year of inclusion into the trial. They were formed based on geographical proximity, with Clusters 1, 3 and 6 comprising two Geriatric rehabilitation Centres (GR Centres), Cluster 7 and 8 containing the hospital setting, and the remaining clusters containing one GR Centre (see Table 1).

**Table 1:** Design of the OTHER two-armed stepped-wedge trial.

| Clusters of GR centres and hospitals | Steps (months) |  |  |  |  |  |  |  |  |
| --- | --- | --- | --- | --- | --- | --- | --- | --- | --- |
|  | 1-3 | 3-6 | 6-9 | 9-12 | 12-15 | 15-18** | 18-21** | 21-24** | 24-27** |
| 1. Location Limburg; 2 GR centres | C | OTHER | OTHER | OTHER | OTHER | OTHER | OTHER | OTHER | OTHER |
| 2. Location Noord-Holland; 1 GR centre | C | C | OTHER | OTHER | OTHER | OTHER | OTHER | OTHER | OTHER |
| 3 Location Noord-Holland; 2 GR centres | C | C | C | OTHER | OTHER | OTHER | OTHER | OTHER | OTHER |
| 4 Location Noord-Brabant; 1 GR centre | C | C | C | C | OTHER | OTHER | OTHER | OTHER | OTHER |
| 5 Location Gelderland: 1 GR centre | C | C | C | C | C | OTHER | OTHER | OTHER | OTHER |
| 6** Location Utrecht; 2 GR centres |  |  |  |  |  | C | C | OTHER | OTHER |
| 7** Location Gelderland; hospital |  |  |  |  |  | C | C | OTHER | OTHER |
| 8 ** Location Amsterdam; hospital |  |  |  |  |  | C | C | C | OTHER |

C= Care as usual, OTHER= OTHER intervention GR = Geriatric Rehabilitation
Trial duration = 18 months (recruitment), 24 months (treatment and measurements) \*\* recruitment extension period of 9 months after evaluation, with 27 months for inclusion.
Pre-rollout period = 3 months. Rollout period = 21 months. Post-rollout period = 6 months.
Step length = 3 months. Average number of participants per step = 6 and 3 (steps 1-5, and 6-7, respectively).

The participating GR centres are Vivium zorggroep (Naarden), GRZPLUS organisation, which is a collaboration with GR centres Omring and Zorgcirkel, Stichting Cicero Zorggroep (Brunssum), Stichting Tante-Louise (Bergen op zoom), Sevagram (Heerlen) and ZZG Herstelcentrum voor revalidatie en zorg (Groesbeek).

After one year of inclusion, the inclusion rate of six persons post stroke (further called ‘the client participants’) every three months per cluster was not reached. We therefore adjusted the design by adding two additional clusters: one cluster containing two GR centres; treatment Centre SZR (Tiel) and AxionContinu (Utrecht) and two clusters each containing one hospital Onze Lieve Vrouwe Gasthuis (OLVG) (Amsterdam), Amsterdam Universitair Medisch Centrum and Canisius-Wilhelmina hospital (Nijmegen). We expect two to three participants per cluster every three months, starting one year after the date of inclusion. Table 1 presents the designs and their corresponding clusters.

Baseline data will be collected within seven days after inclusion of a participant into the trial, 4, 13, and 26 weeks after discharge from clinical rehabilitation or from the hospital. Figure 1 shows the study flow for participants.

**Fig 1:**
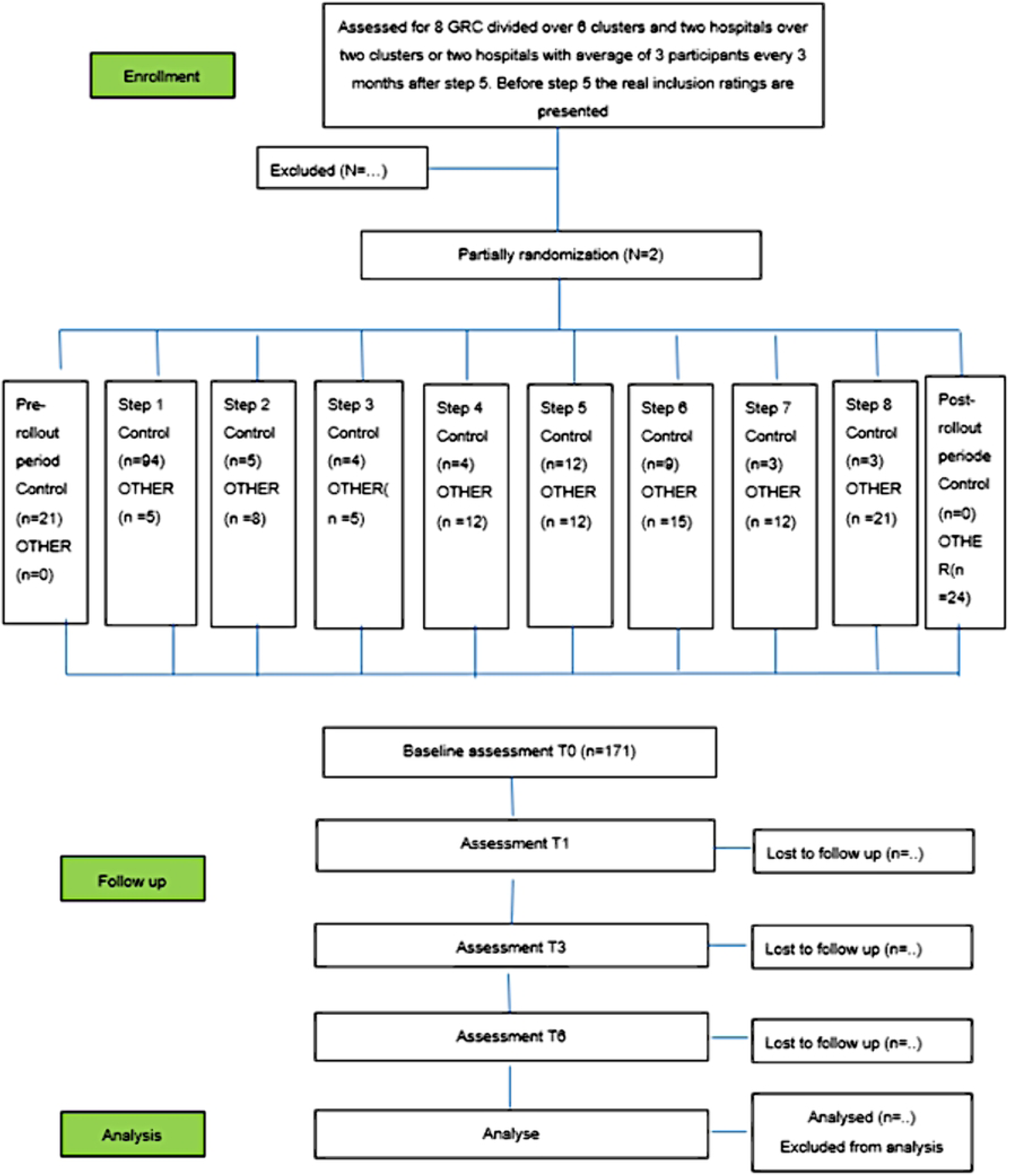
Study flow of clusters and participants of the OTHER trial.

### Participants

The participants we will include in this study are persons post-stroke admitted to one of the nine participating GR centres who are starting inpatient rehabilitation to return home (see Table 1) or those discharged from the participating hospitals (see Table 1). The persons post-stroke are eligible if: 1) they can walk at least a few steps with or without a walking device, 2) are 60 years or older, 3) have a score of at least 16 on the Montreal Cognitive Assessment (MoCA) and 4) have an indication for follow-up home-based GR. Persons post-stroke are ineligible if: 1) they are terminally ill, or 2) they cannot understand verbal information (usually due to severe aphasia).

### Process evaluation

The process evaluation will be conducted using mixed methods based on the MRC framework guidance for process evaluation(24). Quantitative measurement will be used for 1) treatment fidelity (intervention dose, protocol process adherence) by filling in a logbook after every coaching session by OTHER; 2) level of treatment self-reported enactment of participants; 3) participants’ satisfaction with OTHER and 4) Implementation of the intervention (training of occupational therapists, manual for occupational therapists and practical application). The professionals will record details of the intervention dose in the case notes, and data for protocol process adherence by the professionals will be extracted from a structured list in the case notes.

Qualitative data generation: individual semi-structured interviews with a randomly invited number of persons post-stroke; if the partner or informal caregiver was involved during the OTHER-intervention, they will be asked to join the interview (N=15) from starting in February 2025 until December 2025. Focus groups will be used to evaluate OTHER from the professionals’ perspectives on delivering OTHER (N= 8-10). The focus groups are planned in November 2025. The process evaluation will provide insight into the process of implementing OTHER when continuing rehabilitation at home during this trial, as well as how persons post-stroke and professionals reflect on how OTHER supports application in daily life.

## Interventions

The components of the usual care group, the intervention group starting in the GR centre, and the intervention group of people admitted from the hospital to home care are described below.

### Care as usual for participants post-stroke in the geriatric care centre

Inpatient GR begins with an intake, which may be conducted by the geriatric centre’s elderly care physician alone or as part of a multidisciplinary team. After the intake, assessments are conducted, followed by treatment from the multidisciplinary team. Most teams consist of an elderly care physician (a physician specially trained in the medical care of frail or older patients), a nurse, a physical therapist, and an occupational therapist. Other allied health professionals, such as dieticians, social workers or psychologists, are consulted on demand. The elderly care physician coordinates the participant’s multidisciplinary care and treatment plan.

After assessments and observations, a multidisciplinary care and treatment plan is designed in the first multidisciplinary meeting, which is discussed with the person post-stroke and their primary caregiver(s). All persons post-stroke follow a personalised, multidisciplinary rehabilitation program, adjusted to their goals and preferences. In this multidisciplinary meeting, a preliminary indication for home-based GR is made. Home-based GR depends on the person’s needs post-stroke and how care is organised in the area where the person lives. Home-based GR can include allied health professionals, such as occupational therapists, physical therapists and, dietitians, and can also include nurses, social workers or psychologists. Occupational therapy is sometimes involved in home-based GR, but not for all persons post-stroke and not for all organisations. Persons post-stroke are discharged home if they can safely function at home, either independently or with (in)formal care assistance.

### OTHER intervention

When persons post-stroke are included for OTHER, they will receive the OTHER intervention after a short period of inpatient rehabilitation in a GR centre or after discharge from the hospital to their homes. The OTHER intervention is an intervention combining activity monitoring, coaching at the person’s home, and hybrid coaching. The intervention is based on the Self-Determination Theory (26,27), on principles of solution-focused brief therapy (28–31) and occupational performance coaching (32,33). Self-Determination Theory highlights the importance of autonomy, competence, and relatedness, fostering intrinsic motivation by enabling participants to set meaningful goals and exercise control over their rehabilitation process (26,27). Solution-Focused Brief Therapy adopts a strengths-based perspective that prioritises solutions over problems, encouraging individuals to recognise progress and build on their existing capacities (28–31). Occupational Performance Coaching is a person-centred approach that integrates emotional support, structured goal-setting, and knowledge sharing to enhance individuals’ performance, self-efficacy, and problem-solving skills in performing meaningful daily activities (32,33). Collectively, these theoretical frameworks ensure that the intervention was person-centred, empowering participants to take an active role in their rehabilitation and sustain engagement in everyday life. Collaborating with the persons post-stroke is fundamental throughout the intervention and constitutes a core component of Occupational Performance Coaching (28–31). The activity monitoring utilises a validated digital platform, and coaching is conducted either face-to-face or online via videoconferencing.

The intervention for persons post-stroke in GR is intended to start before discharge (at least three weeks before discharge) and with a follow-up at home during 12 weeks. Persons post-stroke who are discharged from the hospital will start as soon as possible with the intervention when they are at home and for 12 weeks.

An occupational therapist who delivers the OTHER intervention will coach persons post-stroke during rehabilitation post-discharge at home to improve daily functioning. During the coaching by the OTs in OTHER, the person post-stroke will be empowered in self-management and improving health-related quality of life.

#### Description of the technology used in OTHER

The activity monitoring intervention tool (Hipper https://hippertx.nl) includes a wearable physical activity monitor (PAM AM300) (https://pamcoach.com) and a gateway (Raspberry Pi). The PAM consists of a 3-dimensional accelerometer, is worn on the hip, and measures 1) the amount of all daily activities in minutes per day and 2) the acceleration of body movements. The measured movements are expressed in a PAM-score representing the energy ratio expended through physical activity compared to resting energy. The PAM communicates with the gateway placed in the person’s home via a Bluetooth adaptor (WR300-E). Via a web application, users can view visualisations of their data on a tablet, phone, or computer.

Once it is clear that a person post-stroke is going home, the person will start wearing a PAM during the inpatient rehabilitation during the day (in the morning from waking up until they go to sleep at night). Through this introduction to activity monitoring, the person learns, with the help of the occupational therapist or a nurse, to wear the sensor every day. The therapist and person post-stroke monitor the activities via a secure web application. The therapist uses the data as feedback to coach the person once a week during a coaching session. In Tables 2 and 3, the components of the intervention are described from the GR centre (Table 2) and from the hospital (Table 3).

**Table 2.** Components of the care as usual and of OTHER delivered from GR setting to persons post-stroke.

| During care in GR Centre | Time frame | Intervention component | Professional involved | Care as usual | OTHER |
| --- | --- | --- | --- | --- | --- |
|  | Multidisciplinary meeting (usually after 2 weeks, sometimes earlier) | Geriatric and multidisciplinary assessment | Elderly care physician, Occupational therapist, and other team members | X | X |
|  |  | Preliminary care and treatment plan |  |  |  |
|  | During inpatient GR | Multidisciplinary rehabilitation | Multidisciplinary team | X | X |
|  | During inpatient GR | Information and instructions on wearing the activity sensor | Occupational therapist or Nurse |  | X |
|  | During inpatient GR | Wearing the activity sensor before discharge | Occupational therapist |  | X |
|  | During inpatient GR | Once a week coaching with the use of sensor data, following the coaching steps | Occupational therapist |  | X |
|  | Last week, before discharge home | Instructions for installing the sensor at home | Occupational therapist, partner |  | X |
| **At Home** | After discharge from inpatient GR centre | Installing the sensor system and wearing the activity monitor | Person post-stroke/ partner/family |  | X |
|  | Week 1 | OTHER and practicing video conferencing | Occupational therapist |  | X |
|  | Week 2 | OTHER * | Occupational therapist |  | X |
|  | Week 3 | OTHER * | Occupational therapist |  | X |
|  | Week 4 | OTHER * | Occupational therapist |  | X |
|  | Weeks 5, 6, 7, 8, 10 | OTHER * | Occupational therapist |  | X |
|  | Week 12 | Removal of the system | Research assistant or Occupational therapist |  | X |

\* Videoconferencing can be used during these consults in dialogue with the participant. Advice is to use videoconferencing 4 times when a participant is at home. GR= Geriatric rehabilitation; OTHER= OTHER intervention

**Table 3.** Components of the care as usual and of OTHER delivered from the hospital setting to persons post-stroke.

| During care in Hospital | Time frame | Intervention component | Professional involved | Care as usual | OTHER |
| --- | --- | --- | --- | --- | --- |
|  | occupational therapist in the hospital | Assessments and to assess going home, GR, or somewhere else | Occupational therapist | X | X |
|  | occupational therapist in the hospital or a transfer nurse | Refer to the occupational therapist in primary care who are involved in OTHER | Occupational therapist/ transfer nurse |  | X |
| At Home | After hospital discharge | Installing the sensor system and wearing the activity monitor | Occupational therapist /Person post-stroke/ partner/family |  | X |
|  | Week 1 | OTHER and practicing video conferencing | Occupational therapist |  | X |
|  | Week 2 | OTHER * | Occupational therapist |  | X |
|  | Week 3 | OTHER * | Occupational therapist |  | X |
|  | Week 4 | OTHER * | Occupational therapist |  | X |
|  | Weeks 5, 6, 7, 8, 10 | OTHER * | Occupational therapist |  | X |
|  | Week 12 | Removal of the system | Research assistant or Occupational therapist |  | X |

\* Videoconferencing can be used during these consults in dialogue with the participant. Advice is to use videoconferencing 4 times when a participant is at home. OTHER= OTHER intervention

#### Coaching by occupational therapist in OTHER

The coaching will take place once a week during the rehabilitation period. Physical coaching sessions at home alternate with online coaching. These are based on the five coaching steps (Figure 2). The web application gives the therapist and the person post-stroke access to the collected data through a personal login. The issues and priorities that are related to daily activities and which are relevant and essential to the person are the point of departure of the coaching sessions. The sensor data can be used to discuss current activity levels performed daily or weekly.

**Fig 2:**
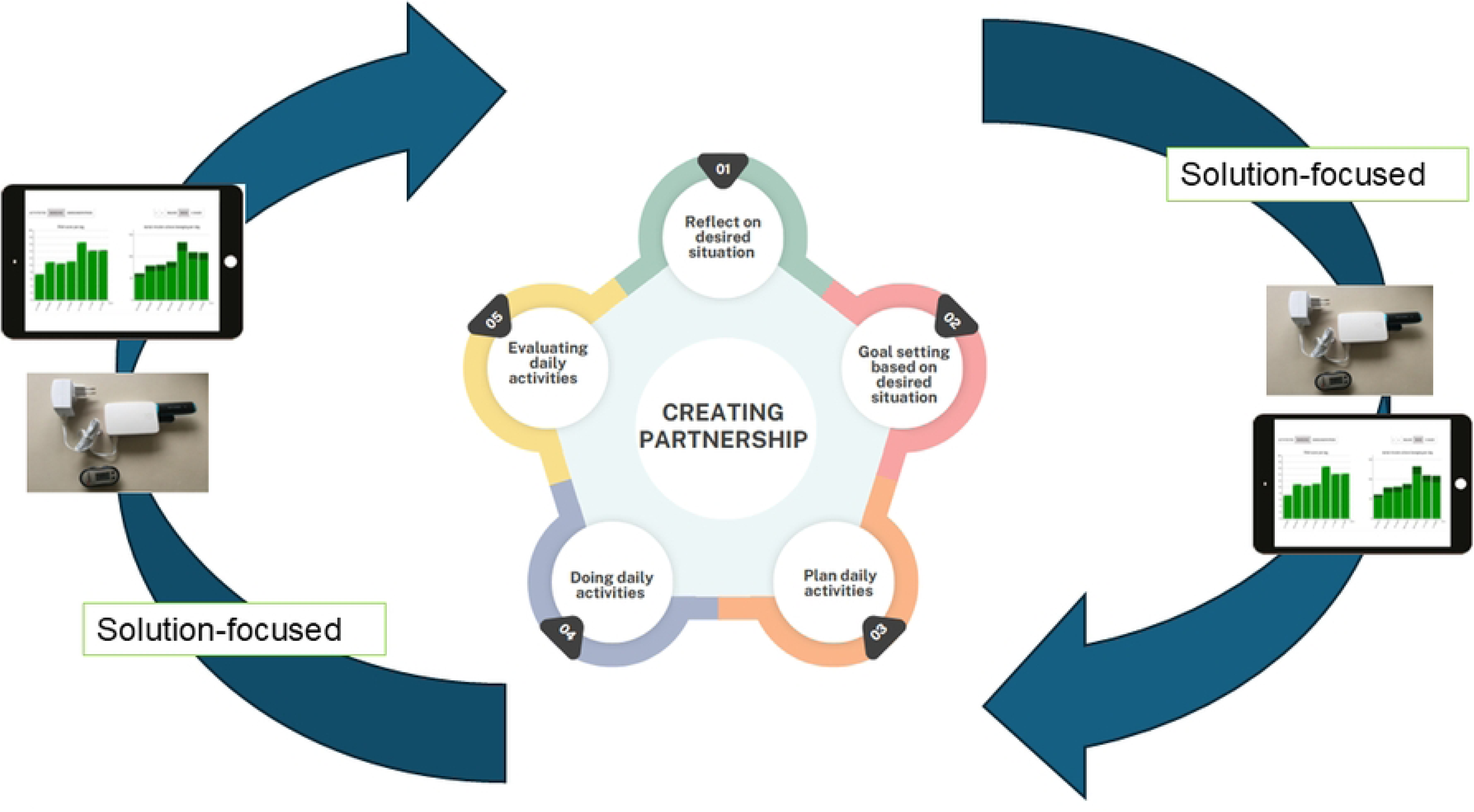
The five coaching steps of the OTHER intervention for the occupational therapist and the use of activity monitoring.

The focus of the coaching is on helping persons post-stroke think about possibilities and take small steps to increase their daily activities.

The five coaching steps are:

The basis of this coaching exists in creating a partnership for working together on an equal footing. According to the theory, this basis is necessary in every coaching step to make coaching effective and optimally support self-management. This basis is a principle of occupational performance coaching (32) and self-determination theory (26,34), with step 1 also being based on solution-focused brief therapy (28–30).

<u>Step 1: Reflection on the desired situation:</u> on daily activities that are important for persons post-stroke and the person’s overall activity during the day, and how this affects their well-being.

<u>Step 2: Goal setting based on the desired situation:</u> the occupational therapist supports the person post-stroke in setting goals that represent daily activities and participation that are important and meaningful for the person post-stroke.

<u>Step 3: Plan daily activities</u>: Plan daily activities by translating the goals and daily activities into small and achievable steps. Also, a coping plan is made to address barriers and facilitators and act on these. Working on small steps is based on solution-focused brief therapy (28–30).

<u>Step 4: Doing activities</u>: The person post-stroke will perform daily activities to see what works and take small steps without the occupational therapist. The occupational therapist will support the person; daily activities will be discussed and evaluated. The person can do the activities alone or with support from the occupational therapist if necessary.

<u>Step 5: Evaluating daily activities:</u> This step evaluates the performance of daily activities, self-management in daily life, the person’s activity level in general throughout the day, and appointments for the next (small) steps.

The sensor data reports are a starting point for the conversation about the daily patterns and activities essential to practice and making new and, realistic plans for following activities based on these objective reports and the reflections on the person’s experiences. The daily and weekly reports can also be used to evaluate the progress of the rehabilitation.

In addition to coaching at the participants’ homes, online coaching sessions will support persons post-stroke. Occupational therapists will use the software available at the GR organisation to book and start a videoconference. The Hipper system will be available for persons post-stroke during this trial.

#### Training and education of the occupational therapists

Every three months, the occupational therapists in a cluster that is about to switch to the OTHER intervention will receive the OTHER training, and after completing this training and new inclusion, they will start treating persons post-stroke to the OTHER intervention. The OTHER training consists of a two-day course followed by a one-day refresher. During the course, all occupational therapists will receive information about the study, the procedures, and the coaching in six steps, as well as guidance on how to utilise the sensor for instructing and coaching persons post-stroke (both face-to-face and videoconferencing). In preparation for the course, professionals receive a manual with this information and are trained on the technical aspects of the activity sensor and its use with the web application. Lastly, the professionals will receive training on the trial’s protocol procedures and participant recruitment for this trial.

After the feasibility study (Pellegrom et al, Manuscript submitted for publication), adjustments were made to the occupational therapist’s training to explain the concept of ‘OTHER’ more clearly for participants, to practice using sensor data and coaching techniques more effectively, and to plan consultations after the training.

## Outcomes

### Primary outcome

The primary outcome measure is ‘daily performance’, 4 weeks, 12 weeks and 24 weeks after the start of OTHER compared to baseline functioning, measured with the performance scale of the Canadian Occupational Performance Measure (COPM) (35) The COPM is a person-centred, occupation-focused outcome measure for detecting changes in perceived daily performance over time. The COPM results in a performance score (COPM-p) and a satisfaction score (COPM-s). Research has shown that the COPM exhibits excellent test-retest reliability and measures changes in the performance of daily activities (35). Through a semi-structured interview, persons will prioritise up to five daily activities that are deemed most important and rate each on a 10-point scale regarding perceived performance (COPM-p) (1 = unable to do at all and 10 = able to do exceptionally well). The mean COPM-p will be obtained by dividing the ratings by the number of prioritised activities. Change in scores can be calculated after a reassessment interval to measure the change in the perception of daily performance. A 1.3-point difference between pre- and post-measurement indicates a minimally clinically important difference (35–37). A trained research assistant will complete the COPM interview and score the results in this study.

### Secondary outcome

– *Persons’ post-stroke satisfaction in performing daily functioning* will be measured with the COPM- s (35). Next to the COPM-p, participants rated the prioritised daily activities on a 10-point scale regarding performance satisfaction (COPM-s) (1 = not satisfied at all and 10 = extremely satisfied). The mean COPM-s will be obtained by summing the ratings and dividing them by the number of prioritised activities. The change scores can be calculated as described above.
– *Self-management.* The Patient-Reported Outcome Measure in Occupational Therapy (PROM-OT) (38,39) is developed to measure the outcome and quality of occupational therapist in the Netherlands from a client’s perspective. The PROM-OT contains 13 questions regarding the outcome of occupational therapist for the person (e.g., I can perform my daily activities, whether or not with help or devices, such as self-care, household, leisure, and work), using a 10-point scale for scoring. It focuses on daily activities, being able to self-manage related to daily functioning and satisfaction of occupational therapist. Key dimensions of self-management in this questionnaire were used to assess self-management, including cognitive understanding of disease-related limitations (Q4), help-seeking behaviour (Q5), boundary setting (Q6), energy management (Q7), emotion acceptance (Q8), and practical problem solving (Q9).
– *Health-related quality of life.* The EuroQol-5D-5L(40) will be used to measure ‘health-related quality of life’ (HRQOL). The EQ-5D-5L measures a patient’s health state using five health dimensions: mobility, self-care, usual activities, pain/discomfort and anxiety/depression, with five severity levels: no problems, slight problems, moderate problems, severe problems and extreme problems. For the economic evaluation, the patients’ EQ-5D-5L health states will be converted into utility scores using the Dutch tariff (41), after which Quality Adjusted Life Years (QALYs) will be estimated using the “Area Under the Curve” approach (42).
– *Capability of older people.* The ICEpop CAPability measure for Older people (ICECAP-O) is a measure of capability in older people for use in economic evaluation(43). Unlike most profile measures used in economic evaluations, the ICECAP-O focuses on wellbeing, which is defined in a broader sense, rather than health.
– *Activity level.* The PAM will be applied to measure the amount of active movement in minutes per day (see description of the Hipper tool) (44,45).
– *Mobility functioning.* Timed Up and Go test (TUG)(46) is a screening tool used to test basic mobility skills of frail elderly patients (60-90 years old) and persons with stroke. The TUG is a general physical performance test used to assess mobility, balance and locomotor performance in elderly people with balance disturbances(47).
– *Satisfaction with the occupational therapist.* The Patient-Reported Outcome Measure – Occupational Therapy (PROM-OT)(38) is developed to measure the outcome and quality of occupational therapist in the Netherlands from a client perspective. The PROM-OT contains 13 questions regarding the outcome of occupational therapist for the person (e.g. I can perform my daily activities, whether or not with help/devices (for example, self-care, household, leisure, work). For information about, see the previous ‘self-management’ parameter description.
– *Societal costs.* Costs will be measured from a societal perspective. This means that all costs related to the intervention (usual care and OTHER) are measured and valued, irrespective of who pays for them or benefits from them. Societal costs will include intervention costs, other healthcare costs (i.e. primary, secondary, complementary, and medication costs), as well as the costs of informal care, unpaid productivity losses, and paid productivity losses. The latter includes both the cost of absenteeism (i.e., sick leave) and presenteeism (i.e., reduced productivity while at work). Intervention costs will be assessed through micro-costing, meaning that detailed data will be collected on resource consumption during the intervention and their respective unit prices. For all other cost categories, resource consumption will be measured using iMCQ-, iPCQ-, and iVICQ-based retrospective cost questionnaires administered after 13 and 26 weeks. Healthcare costs will be valued using Dutch standard costs and prices derived from www.medicijnkosten.nl. Informal care (i.e. care by family and friends) and unpaid productivity losses (i.e. costs associated with reduced productivity levels related to unpaid activities, such as volunteer work) will be valued using a recommended Dutch shadow price. Absenteeism will be valued according to the Friction Cost Approach and using gender-specific price weights. Presenteeism will be valued using gender-specific price weights as well (48).

Besides the outcomes mentioned above, at baseline, data will be collected on:

– *Sociodemographic characteristics*: age, gender, date and time of admission to hospital or geriatric clinic, date of discharge, the highest level of education, marital status, and living arrangement.
– *Chronic conditions* will be measured by the Functional Comorbidity Index (FCI) (49). The FCI is a sum of 18 self-reported comorbid conditions with a score of 0 to 18. A score of 0 indicates no comorbid illness, and a score of 18 indicates the highest number of comorbid illnesses.
– *Cognitive functioning*. To classify the severity of cognitive impairment, the Montreal Cognitive Assessment (MoCA)(50) will be used. The MoCA is a 30-point test that can be administered in 10 minutes.

### Process evaluation

The process evaluation will be conducted using a mixed-methods approach, and the process evaluation of complex interventions from the Medical Research Council guidance will be used (24). Quantitative measurement will be used for 1) treatment fidelity (intervention dose, protocol process adherence) by occupational therapists filling in a logbook after every intervention by OTHER; 2) level of treatment enactment of persons post stroke from individual interviews and questionnaire during measurements at home; 3) participants’ satisfaction with OTHER by a questionnaire during the measurements at home (one month, three months and six months after discharge); and 4) Implementation of the intervention (training of OT, manual for OT’s and practical application). Details on intervention dose will be recorded by the occupational therapist in the case notes, and data for protocol process adherence by the professionals will be extracted from a structured list in the case notes, see Table 4.

**Table 4:** Variables and outcome measures and time points of assessment in.

|  | Enrolment |  |  |  |  |
| --- | --- | --- | --- | --- | --- |
| Outcome measure | Instrument or Question | Baseline | T1 | T2 | T3 |
| **Primary outcome measure** |  |  |  |  |  |
| Perceived daily functioning – self-perceived performance in daily activities | COPM | X | X | X | X |
| **Secondary outcome measure** |  |  |  |  |  |
| Psychosocial functioning: |  |  |  |  |  |
| Self-management | PROM- OT | X | X | X | X |
| Quality of life | EQ-5D-5L | X | X | X | X |
| Capability of older people | ICECAP-O | X | X | X | X |
| Activity and mobility functioning: |  |  |  |  |  |
| Activity level | PAM | X | X | X | X |
| Mobility functioning | TUG | X | X |  | X |
| Satisfaction of occupational therapist: |  |  |  |  |  |
| Satisfaction of occupational therapist: | PROM-OT | X | X | X | X |
| Health care utilization/ Societal cost |  |  |  |  |  |
| Consult general practitioner/ PT/ dietician etc. | CQ |  |  | X | X |
| Consult specialist | CQ |  |  | X | X |
| Alternative care | CQ |  |  | X | X |
| Care in house | CQ |  |  | X | X |
| Assistive devices | CQ |  |  | X | X |
| Medication use | CQ |  |  | X | X |
| Work | CQ |  |  | X | X |
| Caregiver | CQ |  |  | X | X |
| **Additional measures:** Information gathered on the determinants of functional decline (e.g., comorbidities) and a minimal data set (MDS) consisting of; |  |  |  |  |  |
| Demographic data |  | X |  |  |  |
| Medical comorbidity - FCI |  | X |  |  | X |
| Cognitive functioning – MoCA | X |  |  |  | X |
| Process evaluation - Treatment Fidelity |  |  |  |  |  |
| Logfile completed | Logbook OT | X | X | X |  |
| Intervention dose - Number sessions delivered | Logbook OT |  | X | X |  |
| Protocol process adherence- Once a week coaching with the sensor data during GRC and at home | Logbook OT |  |  |  |  |
| Protocol process adherence - Case conference during home-based rehabilitation | Logbook OT and PQ |  | X | X |  |
| Process evaluation- Level of treatment enactment participant- Daily wearing of the activity sensor<br>Y or no. If no reason | Logbook OT, PQ and sensor database |  | X | X |  |
| Once a week looking at the sensor data with therapist<br>Y or no. If no reason | Logbook OT and PQ |  | X | X |  |
| Videoconferencing<br>Y or no. if no reason | Logbook OT and PQ |  | X | X |  |
| Process evaluation- Participants' experiences with OTHER | Individual interview |  |  |  | X |
| Process evaluation- Occupational therapists experience with OTHER | Focus group |  |  |  | X |

B= Baseline. T1= 4 weeks after discharge (home visit). T2= 13 weeks after discharge (home visit). T3= 26 weeks after discharge (home visit). COPM= Canadian Occupational Performance Measure-performance measure. PROM-OT= Patient-Reported Outcome Occupational Therapy. EQ5D= Euro QoL-5D-L. ICECAP-O= ICEpop CAPability measure for Older people. PAM= Physical activity monitoring. TUG= Time up and go test. MoCA= Montreal Cognitive assessment. PROM-OT= Patient-Reported Outcome Measure – Occupational Therapy. PAM= physical Activity Measure. CQ= Care questionnaire FCI= Functional Comorbidity Index. PQ= Process Questionnaire for person post-stroke

Qualitative data generation: individual semi-structured interview with a randomly invited number of persons post-stroke; if a partner or caregiver was involved during the OTHER-intervention, they will be asked to join the interview (N=15). Topics of the interview will include satisfaction with the intervention, content, delivery, results of rehabilitation on daily functioning and participation in joint goal setting, tailoring of the rehabilitation, as well as barriers and facilitators. A focus group will be conducted to evaluate OTHER from the professionals’ perspectives on delivering OTHER, the mechanism of impact, contextual factors that influence the intervention, barriers and facilitators of implementing OTHER during this trial (N=8-10).

Table 4 gives a detailed overview of outcome measures at each time point.

### Data collection methods

Trained research assistants will provide information about the research, obtain informed consent for people after stroke and informal caregivers and all outcome assessments for the GR centre: at baseline, 4 weeks home (T1), 12 weeks at home (T2) and a follow-up 26 weeks at home (T3). Measurements for the effect are taken at baseline in GR centre, and subsequent measurements are conducted at home. For the cost questionnaire, a second phone call appointment will be made to complete the questions. The feasibility study made clear that the measurements are exhausting for the participants. For this reason, measurements are split up. Data will be obtained from the electronic patient record (EPR) through standardised questionnaires and assessments. All data will be entered into a database, CASTOR, in accordance with good clinical practice guidelines, using an identification code for each participant.

### Sample size

The initial sample size calculation at the beginning of the recruitment yielded a sample size of 180 based on the following assumptions:

- A 1.19 point between-group difference on the primary outcome (COPM) as the smallest difference of clinical interest,
- a standard deviation of 1.70 (around a mean of 6.81) and 1.75 (around a mean of 8.19) in the control and experimental groups, respectively,
- an intraclass correlation of 0.03,
- an alpha of 0.05 and
- an average cluster size of 5
- a sample size of 180 (i.e. 150 participants in total; 5 participants × 5 steps × 6 periods (since all centres start in the control condition) yielding a power of 0.7998. Accounting for a 20 per cent dropout rate, yielding a required sample size of 180 (19).

A new sample size calculation was performed one year after the start of recruitment because the number of participants included was much lower than we had aimed for. Using the exact same assumptions as above and adding an additional assumption of an average cluster size of 2 or 3 participants in the new GR centres, the sample size went up to 114 (137 assuming 20% loss to follow-up) for the care as usuals group and 171 (205 assuming 20% loss to follow-up) for the OTHER group respectively, with corresponding powers of 0.7843 and 0.9093, respectively. For more information about the STATA command used to calculate sample size, see S 1.

### Randomisation

Figure 1 shows the flow of clusters and participants post-stroke through the trial. We deemed obtaining equal amounts of patient outcome information for each treatment condition as more important than unrestrictedly randomising the five clusters for two reasons. First, randomisation of so few units does not guarantee baseline comparability. Second, and related to the first reason, statistical adjustment of treatment effect estimates will be needed, which is more effective if both treatment groups have comparable sizes. Thus, assignment of clusters to the five initial sequences was performed four weeks before the start of data collection by GtR, a methodologist, not involved in the day-to-day logistics of care delivery in three steps: (i) the five clusters were ranked in order of the numbers of patients we expected them to recruit (based on prior administrative information obtained from the centres); (ii) the two largest clusters were manually put into sequences 1 and 5, respectively; the intermediate-sized cluster was manually put into sequence 3; (iii) we randomly allocated the remaining two (smallest) clusters into sequence 2 and 4 using the sample command in STATA 18.

### Statistical methods

Comparability at the level of clusters and participants immediately after the assignment of the sequences will be assessed to detect potentially important imbalances in prognostic indicators. Descriptive data will be used to determine any time trends of persons’ post-stroke characteristics at recruitment, since patient selection bias is a threat in cluster trials that cannot be blinded for allocation. Except for one secondary outcome, all outcomes will be analysed using mixed linear regression analysis on measurement times 4, 13 and 26 weeks (with baseline values as a covariate). The crude (unadjusted) treatment effect and its effect after adjustment for a set of relevant confounders will be estimated. Additional decisions on relevant confounders (variables that appear unbalanced at baseline and are not in the confounder set) will be made using the 10 per cent change-in-estimate criterion, which works well in linear regression (as opposed to its use in logistic regression) (51). Participating centres will be represented as dummy variables in all analyses. Models will include a random intercept for participants. Random slopes will be fitted to assess the improvement of the model fit. Using information from regularly updated and semi-standardised occupational therapists’ logbooks and repeated measurements of the main time-varying confounders, we will analyse the treatment effect (if any) as a function of treatment fidelity and, if possible, participants’ adherence using structural mean models (52–54). Baseline comparability of participants will be assessed. Baseline assessments will be summarised using interquartile ranges (for continuous and ordinal variables) or percentages (for proportions).

The main analysis focuses on a comparison between the intervention group OTHER and the care as usual concerning the primary outcome, the perceived daily function scores, with scores at T1, T3 and T6 as dependent variables and the baseline score as a covariate. The differences between the two groups will be evaluated using a mixed linear regression analysis. Two-sided 95% confidence intervals will be calculated. An intention-to-treat analysis will be conducted. The most recent versions of SPSS, STATA or R-CRAN will be used to analyse the data.

The treatment effects on the secondary outcomes will be estimated using the same multi- level approach. Survival data (institutionalisation and mortality) will be analysed flexible parametric survival models (55,56). Restricted mean survival models will be used as a sensitivity analysis where appropriate (57–59). Two-sided 95% confidence intervals will be calculated. Following the abovementioned approach, the ordinal outcome (ICECAP-O) will be analysed using mixed-effects ordinal logistic regression analysis. For more information about the Statistical Analysis Plan, see S2.

Cost-effectiveness analyses will be performed according to the intention-to-treat principle. In the main analysis, missing cost and effect data will be imputed using Multivariate Imputation by Chained Equations (MICE) (60). Rubin’s rules will be used to pool the results from the different multiple imputed datasets. Multilevel analyses will be performed to estimate cost and effect differences between the OTHER-intervention and usual care, while adjusting for confounders if necessary. In doing so, the stepped wedge design of the trial will be considered. Incremental Cost-Effectiveness Ratios (ICERs) will be calculated by dividing the differences in costs between groups by the differences in the primary outcome (i.e. COMP-p) and QALYs. Bias-corrected and accelerated bootstrapping with 5000 replications will be used to estimate 95% confidence intervals around the cost differences and statistical uncertainty surrounding the ICERs. Uncertainty surrounding the ICERs will be graphically presented on cost-effectiveness planes. Cost-effectiveness acceptability curves will be estimated, showing the probability that OTHER-intervention is cost-effective compared to usual care for a range of different ceiling ratios, thereby showing decision uncertainty. Sensitivity analyses will be done to assess the robustness of the results (e.g. complete-case analysis, per-protocol analysis).

Process evaluation will be performed based on a mixed methods design. Methods for qualitative analysis follow the constant comparative method. The individual and focus group will be transcribed verbatim and imported into MAXQDA. The researcher will read the data to get familiarised. Two researchers will perform the analysis and will apply open coding techniques derived from constant comparison methods. Open data coding was conducted by the researcher and a part of the transcript was also coded by the second researcher followed by comparison and discussion of the codes in order to reach consensus on the coding procedure and content. Both researchers will identify potential categories among initial codes, and the potential categories will be discussed by members of the research group and further grouped into final, main themes related to the research question. Quantitative data of the process analysis will be descriptive. The Statistical Analysis Plan is available at https://osf.io/cqnrx/ (under construction).

### Ethics and dissemination

The Medical Research Ethics Committee of Amsterdam UMC has approved the study (protocol ID 2022.0786). Written consent is obtained from all participants prior to their inclusion. The research is performed according to the Dutch Medical Research Involving Human Subjects Act and the WMA Declaration of Helsinki (61).

All information collected during the trial will be kept strictly confidential.

Study results will be published in peer-reviewed journals, reported in line with the literature Consolidated Standards of Reporting Trials guidelines for stepped wedge cluster randomised trials (23).

## Discussion

The present partially randomised stepped-wedge trial evaluates the OTHER intervention on effectiveness, cost-effectiveness and process of implementation, comparing it to usual care. OTHER combines activity monitoring with coaching by occupational therapists, providing them with tools to optimise the transition from clinical rehabilitation to home, focusing on daily activity performance and self-management. This transition is found to be challenging for persons post-stroke, and research has shown that persons post-stroke are often inactive and sedentary (12,13). By providing persons post- stroke with insight into their daily activity patterns and the importance of performing these activities, and coaching them to make small steps and decisions on daily activities, we aim to significantly increase their performance and engagement in activities and self-management. We also expect the intervention to be cost-effective compared to care as usual.

Much research has provided insight into the (technical) development of technology/e-health, which can be utilised by professionals during therapy for persons post-stroke (17,18,20,21). On the other hand, professionals often find it challenging to integrate and implement technology into their clinical practice. A scoping review about the patients’ perspective of GR highlights the need for support (physical, psychological, social, and how to cope with limitations), the need for shared decision-making and autonomy, for a stimulating rehabilitation environment and rehabilitation at home (62). With OTHER, we aim to provide occupational therapists with tools to meet the needs of persons post-stroke and integrate technology into their daily practice.

The strengths of this study include the stepped-wedge design for evaluating (cost-) effectiveness, which has several advantages. All participating occupational therapists will be trained in OTHER and can use the different coaching steps and technology during daily practice. During the feasibility study of OTHER, occupational therapists stated that the use of the coaching steps was also useful with other persons than post-stroke (Pellegrom et al, Manuscript submitted for publication). Also, by using a partially randomised stepped-wedge design, we are able to include different organisations of geriatric rehabilitation across the Netherlands that deliver or refer to geriatric home rehabilitation. Including these different organisations gives insight into the effectiveness of OTHER delivered from and implemented in a broad range of organisations that are usually concerned with the care of people post-stroke. However, the two-stepped wedge design also has challenges in analysis and requires complex logistics and planning. We have eight clusters (hospitals and GR centres), with context-specific findings. We deemed obtaining equal amounts of patient outcome information for each treatment condition more important than unrestrictedly randomising the 5 clusters, for two reasons. First, randomisation of so few units does not guarantee baseline comparability. Second, and related to the first reason, statistical adjustment of treatment effect estimates will be needed, which is more effective if both treatment groups have comparable sizes. We will try to express the effect as a function of fidelity. It was challenging to define the treatment fidelity of the OTHER intervention, which formed the basis for the analysis of treatment fidelity. Analysis of fidelity is described in the Statistical Analysis Plan.

Another challenge is recruiting participants; the expected numbers appeared to be unfeasible with the initial clusters after one year of inclusion. After an evaluation and deliberations with the funder, we have made adjustments to the timeline by extending the data collection by nine months and adding three new clusters. Based on other studies with recruitment problems, another adjustment we made is that occupational therapists will be supported by the research assistant in recruiting participants for this trial (63). A motivating video has been created to illustrate why persons post-stroke are participating in the trial. Therefore, some participants who are already participating in the trial were interviewed. This video will help occupational therapists recruit more participants post-stroke and caregivers for this trial. In parallel, we will conduct a process evaluation to assess the implementation of the intervention and identify any variations that may occur during its implementation.

This partially cluster randomised stepped-wedge trial on the (cost-) effectiveness of the OTHER intervention will also provide information that assists people after stroke in returning home to resume and manage daily lives. The findings will lead to an evidence-based intervention for people post-stroke with information on its cost-effectiveness and will give recommendations for implementation. The partially cluster randomised trial has a stepped-wedge design with use of an intention-to-treat analysis, and includes a process and economic evaluation.

With this protocol paper, we strive to be transparent on the process of our research by preregistering our trial, publishing the protocol and SAP and adhering to the FAIR principles.

### Trial status

Recruitment started on April 23, 2023. We completed recruitment for people after stroke at the end of July 2025. Data collection will continue until the end of January 2026. At the time of submission (October 2025), 121 participants had been enrolled. For the process evaluation, professionals for the focus groups (November 2025) are recruited now and individual interview of people post-stroke and informal caregiver started in February 2025 and will go on until December 2025.

## Data Availability

No datasets were generated or analysed during the current study. All relevant data from this study will be made available upon study completion.

## Authors’ contributions

**Conceptualization:** Margriet C. Pol, Ton Satink, Margo van Hartingsveldt, Bianca Buurman, Maud Graff.

**Funding acquisition**: Margriet C. Pol.

**Methodology:** Sanne Pellegrom, Gerben ter Riet, Margriet C. Pol, Johanna Maria van Dongen.

**Project administration**: Sanne Pellegrom, Margriet C. Pol.

**Writing- original draft**: Sanne Pellegrom.

**Writing- review & editing:** Margriet C. Pol, Ton Satink, Margo van Hartingsveldt, Johanna Maria van Dongen, Gerben ter Riet, Bianca M. Buurman, Maud Graff.

## Acknowledgements

We would like to thank all the involved persons post-stroke and their caregivers, The participating GR centres are Vivium zorggroep (Naarden), GRZPLUS organisation is a collaboration with GR centres Omring and Zorgcirkel, Stichting Cicero Zorggroep (Brunssum), Stichting Tante-Louise (Bergen op zoom), Sevagram (Heerlen), ZZG Herstelcentrum voor revalidatie en zorg (Groesbeek), behandelcentrum SZR (Tiel) and AxionContinu (Utrecht). For the hospital: OLVG (Amsterdam), Amsterdam Universitair Medisch Centrum and Canisius-Wilhelmina Ziekenhuis (Nijmegen) and occupational therapists for participating in the OTHER project.

## Supporting Information

S1 Study Protocol

S2 Matrices from the steppedwedge command in STATA

S3 SPIRIT checklist

## References

1. Vat LE, Middelkoop I, Buijck BI, Minkman MMN. The Development of Integrated Stroke Care in the Netherlands a Benchmark Study. Int J Integr Care. 2016 Nov 16;16(4).

2. Feigin VL, Brainin M, Norrving B, Martins S, Sacco RL, Hacke W, et al. World Stroke Organization (WSO): Global Stroke Fact Sheet 2022. International Journal of Stroke [Internet]. 2022 Jan 5;17(1):18–29. Available from: https://journals.sagepub.com/doi/10.1177/17474930211065917

3. Norrving B, Barrick J, Davalos A, Dichgans M, Cordonnier C, Guekht A, et al. Action Plan for Stroke in Europe 2018–2030. Eur Stroke J [Internet]. 2018 Dec 29;3(4):309–36. Available from: https://journals.sagepub.com/doi/10.1177/2396987318808719

4. Li X yu, Kong X meng, Yang C hao, Cheng Z feng, Lv J jie, Guo H, et al. Global, regional, and national burden of ischemic stroke, 1990–2021: an analysis of data from the global burden of disease study 2021. EClinicalMedicine [Internet]. 2024 Sep 1;75:102758. Available from: https://linkinghub.elsevier.com/retrieve/pii/S2589537024003377

5. Satink T, Cup EHC, de Swart BJM, Nijhuis-van der Sanden MWG. How is self-management perceived by community living people after a stroke? A focus group study. Disabil Rehabil [Internet]. 2015 Jan 30;37(3):223–30. Available from: http://www.tandfonline.com/doi/full/10.3109/09638288.2014.918187

6. Quinn K, Murray C, Malone C. Spousal experiences of coping with and adapting to caregiving for a partner who has a stroke: a meta-synthesis of qualitative research. Disabil Rehabil. 2014 Feb 18;36(3):185–98.

7. Jammal M, Kolt GS, Liu KPY, Dennaoui N, George ES. The impact of caregiving on the roles and valued activities of stroke carers: A systematic review of qualitative studies. PLoS One. 2024 May 1;19(5).

8. Winstein CJ, Stein J, Arena R, Bates B, Cherney LR, Cramer SC, et al. Guidelines for Adult Stroke Rehabilitation and Recovery. Stroke. 2016 Jun;47(6).

9. Tol-Schilder M, van Zijl L, Slee-Valentijn M, Wattel L. Flowcard CVA voor geriatrische revalidatiezorg (GR). Tijdschrift voor ouderengeneeskunde [Internet]. 2020;1–12. Available from: https://www.verenso.nl/magazine-augustus-2020/no-4-augustus-2020/wetenschap/flowcard-cva-voor-geriatrische-revalidatiezorg-gr

10. Nederlands Zorgautoriteit. Open data of Nederlands Zorgautoriteit [Internet]. [cited 2025 Apr 10]. Available from: https://www.opendisdata.nl/

11. Ezeugwu VE, Garga N, Manns PJ. Reducing sedentary behaviour after stroke: perspectives of ambulatory individuals with stroke. Disabil Rehabil. 2017 Dec 4;39(25):2551–8.

12. Wondergem R, Veenhof C, Wouters EMJ, de Bie RA, Visser-Meily JMA, Pisters MF. Movement Behavior Patterns in People With First-Ever Stroke. Stroke [Internet]. 2019 Dec;50(12):3553–60. Available from: https://www.ahajournals.org/doi/10.1161/STROKEAHA.119.027013

13. Kleindorfer DO, Towfighi A, Chaturvedi S, Cockroft KM, Gutierrez J, Lombardi-Hill D, et al. 2021 Guideline for the Prevention of Stroke in Patients With Stroke and Transient Ischemic Attack: A Guideline From the American Heart Association/American Stroke Association. Vol. 52, Stroke. Wolters Kluwer Health; 2021. p. E364–467.

14. Zonneveld M, Patomella AH, Asaba E, Guidetti S. The use of information and communication technology in healthcare to improve participation in everyday life: a scoping review. Disabil Rehabil [Internet]. 2020 Nov 5;42(23):3416–23. Available from: https://www.tandfonline.com/doi/full/10.1080/09638288.2019.1592246

15. Cogollor JM, Rojo-Lacal J, Hermsdörfer J, Ferre M, Arredondo Waldmeyer MT, Giachritsis C, et al. Evolution of Cognitive Rehabilitation After Stroke From Traditional Techniques to Smart and Personalized Home-Based Information and Communication Technology Systems: Literature Review. JMIR Rehabil Assist Technol [Internet]. 2018 Mar 26;5(1):e4. Available from: http://rehab.jmir.org/2018/1/e4/

16. Zonneveld M, Patomella AH, Asaba E, Guidetti S. The use of information and communication technology in healthcare to improve participation in everyday life: a scoping review. Disabil Rehabil [Internet]. 2020;42(23):3416–23. Available from: 10.1080/09638288.2019.1592246

17. Nam HS, Park E, Heo JH. Facilitating Stroke Management using Modern Information Technology. J Stroke. 2013;15(3):135.

18. Dumitrascu OM, Demaerschalk BM. Telestroke. Curr Cardiol Rep. 2017 Sep 7;19(9):85.

19. Ware LJ, Rennie KL, Schutte AE. Monitoring physical activity after a cardiovascular event: What is ‘fit’ for purpose? Vol. 25, European Journal of Preventive Cardiology. SAGE Publications Inc.; 2018. p. 220–2.

20. Hestetun-Mandrup AM, Toh ZA, Oh HX, He HG, Martinsen ACT, Pikkarainen M. Effectiveness of digital home rehabilitation and supervision for stroke survivors: A systematic review and meta-analysis. Digit Health. 2024;10.

21. Westlake K, Akinlosotu R, Udo J, Goldstein Shipper A, Waller SMC, Whitall J. Some home- based self-managed rehabilitation interventions can improve arm activity after stroke: A systematic review and narrative synthesis. Front Neurol. 2023;14.

22. Pol MC, ter Riet G, van Hartingsveldt M, Kröse B, Buurman BM. Effectiveness of sensor monitoring in a rehabilitation programme for older patients after hip fracture: a three-arm stepped wedge randomised trial. Age Ageing [Internet]. 2019 Sep 1;48(5):650–7. Available from: https://academic.oup.com/ageing/article/48/5/650/5519556

23. Hemming K, Taljaard M, Grimshaw J. Introducing the new CONSORT extension for stepped- wedge cluster randomised trials. Vol. 20, Trials. BioMed Central Ltd; 2019.

24. Moore GF, Audrey S, Barker M, Bond L, Bonell C, Hardeman W, et al. Process evaluation of complex interventions: Medical Research Council guidance. BMJ (Online). 2015 Mar 19;350.

25. Skivington K, Matthews L, Simpson SA, Craig P, Baird J, Blazeby JM, et al. A new framework for developing and evaluating complex interventions: Update of Medical Research Council guidance. The BMJ. 2021 Sep 30;374.

26. Ryan RM, Deci EL. Intrinsic and extrinsic motivation from a self-determination theory perspective: Definitions, theory, practices, and future directions. Contemp Educ Psychol. 2020 Apr 1;61.

27. Flannery M. Self-Determination Theory: Intrinsic Motivation and Behavioral Change. Oncol Nurs Forum. 2017 Mar 1;155–6.

28. van Beek Y, Hessen D, Levelt L, Beijer D, Rijnberk C, Maras A, et al. Intensive specialised multi- family therapy for multi-stressed families: Therapeutic alliance as predictor for effectiveness. J Fam Ther. 2023 Aug 1;45(3):271–90.

29. Gan C. Solution-Focused Brief Therapy (SFBT) with individuals with brain injury and their families. NeuroRehabilitation. 2020;46(2):143–55.

30. Klaver M, Bannik F. Oplossingsgerichte therapie bij patiënten met niet-aangeboren hersenletsel. 2010;2(5):11–9.

31. Vermeulen-Oskam E, Franklin C, van’t Hof LPM, Stams GJJM, van Vugt ES, Assink M, et al. The current evidence of solution-focused brief therapy: A meta-analysis of psychosocial outcomes and moderating factors. Clin Psychol Rev. 2024 Dec;114:102512.

32. Graham F, Kennedy-Behr A, Ziviani J. Occupational Performance Coaching. 1st ed. Abingdon, Oxon; New York, NY : Routledge, 2020.: Routledge; 2020.

33. Kessler D, Anderson ND, Dawson DR. Occupational performance coaching for stroke survivors delivered via telerehabilitation using a single-case experimental design. British Journal of Occupational Therapy. 2021 Aug 9;84(8):488–96.

34. Bakker JM, Bannink FP. Solution focused brief therapy in psychiatric practice. Tijdschr Psychiatr [Internet]. 2008 [cited 2025 Apr 15];50(1):55–9. Available from: https://positievegezondheidszorg.nl/site/wp-content/uploads/Oplossingsgerichte_therapie_in_de_psychiatrische_praktijk.pdf

35. Law M, Baptiste SAC, McColl MA, Polatajko HJ PN. Canadian Occupational Performance Measure (COPM) 5th ed. 5th ed. Ottawa: CAOT Publications ACE; 2014.

36. Eyssen ICJM, Steultjens MPM, Oud TAM, Bolt EM, Maasdam A, Dekker J. Responsiveness of the Canadian Occupational Performance Measure. The Journal of Rehabilitation Research and Development. 2011;48(5):517.

37. Sturkenboom IHWM, Graff MJL, Hendriks JCM, Veenhuizen Y, Munneke M, Bloem BR, et al. Efficacy of occupational therapy for patients with Parkinson’s disease: A randomised controlled trial. Lancet Neurol [Internet]. 2014;13(6):557–66. Available from: 10.1016/S1474-4422(14)70055-9

38. van de Ven-Stevens L, Cup E, Wassink D, Satink T, Graff M. Meten is weten, Steeds meer aandacht voor meten vanuit het perspectief van de cliënt: de PRO-Ergo. 3. 2022;14–6.

39. Arnoldus E, Bekkers E, van Dijk S, Hermans Y, Nijland A, Peters M. De validiteit van de PRO- Ergo. Ergotherapie Magazine. 2022;40–7.

40. EuroQol. EuroQol - a new facility for the measurement of health-related quality of life. Health Policy (New York). 1990 Dec;16(3):199–208.

41. M. Versteegh M, M. Vermeulen K, M. A. A. Evers S, de Wit GA, Prenger R, A. Stolk E. Dutch Tariff for the Five-Level Version of EQ-5D. Value in Health. 2016 Jun;19(4):343–52.

42. Drummond MF, Sculpher MJ, Claxton K, Stoddart GL, Torrance GW. Methods for the Economic Evaluation of Health Care Programmes. 4th ed. Oxford: Oxford University Press; 2015. 1–445 p.

43. Proud L, McLoughlin C, Kinghorn P. ICECAP-O, the current state of play: a systematic review of studies reporting the psychometric properties and use of the instrument over the decade since its publication. Qual Life Res. 2019 Jun;28(6):1429–39.

44. De Greef M. The accuracy of the Pam accelerometer in assessing daily physical activity [Internet]. 2014. Available from: https://www.researchgate.net/publication/254695470

45. Slootmaker SM, Chin A Paw MJM, Schuit AJ, van Mechelen W, Koppes LLJ. Concurrent validity of the PAM accelerometer relative to the MTI Actigraph using oxygen consumption as a reference. Scand J Med Sci Sports. 2009 Feb;19(1):36–43.

46. Podsiadlo D, Richardson S. The timed ‘Up & Go’: a test of basic functional mobility for frail elderly persons. J Am Geriatr Soc. 1991 Feb;39(2):142–8.

47. Ng SS, Hui-Chan CW. The timed up & go test: its reliability and association with lower-limb impairments and locomotor capacities in people with chronic stroke. Arch Phys Med Rehabil. 2005 Aug;86(8):1641–7.

48. Zorginstituut Nederland. Richtlijn voor het uitvoeren van economische evaluaties in de gezondheidszorg. 29-02-2016 [Internet]. 2016;(november):120. Available from: https://www.ispor.org/PEguidelines/source/NL-Economic_Evaluation_Guidelines.pdf

49. Groll D, To T, Bombardier C, Wright J. The development of a comorbidity index with physical function as the outcome. J Clin Epidemiol. 2005 Jun;58(6):595–602.

50. Sweet L, Van Adel M, Metcalf V, Wright L, Harley A, Leiva R, et al. The Montreal Cognitive Assessment (MoCA) in geriatric rehabilitation: psychometric properties and association with rehabilitation outcomes. Int Psychogeriatr. 2011 Dec;23(10):1582–91.

51. Schuster NA, Twisk JWR, ter Riet G, Heymans MW, Rijnhart JJM. Noncollapsibility and its role in quantifying confounding bias in logistic regression. BMC Med Res Methodol [Internet]. 2021 Dec 5;21(1):136. Available from: https://bmcmedresmethodol.biomedcentral.com/articles/10.1186/s12874-021-01316-8

52. Cole SR, Hernán MA, Margolick JB, Cohen MH, Robins JM. Marginal Structural Models for Estimating the Effect of Highly Active Antiretroviral Therapy Initiation on CD4 Cell Count. Am J Epidemiol [Internet]. 2005 Sep 1;162(5):471–8. Available from: http://academic.oup.com/aje/article/162/5/471/82419/Marginal-Structural-Models-for-Estimating-the

53. Hernán MA, Brumback BA, Robins JM. Estimating the causal effect of zidovudine on CD4 count with a marginal structural model for repeated measures. Stat Med [Internet]. 2002 Jun 30;21(12):1689–709. Available from: https://onlinelibrary.wiley.com/doi/10.1002/sim.1144

54. Brumback BA, Hernán MA, Haneuse SJPA, Robins JM. Sensitivity analyses for unmeasured confounding assuming a marginal structural model for repeated measures. Stat Med [Internet]. 2004 Mar 15;23(5):749–67. Available from: https://onlinelibrary.wiley.com/doi/10.1002/sim.1657

55. Syriopoulou E, Mozumder SI, Rutherford MJ, Lambert PC. Robustness of individual and marginal model-based estimates: A sensitivity analysis of flexible parametric models. Cancer Epidemiol [Internet]. 2019 Feb;58:17–24. Available from: https://linkinghub.elsevier.com/retrieve/pii/S187778211830328X

56. Lambert PC, Wilkes SR, Crowther MJ. Flexible parametric modelling of the cause-specific cumulative incidence function. Stat Med [Internet]. 2017 Apr 30;36(9):1429–46. Available from: https://onlinelibrary.wiley.com/doi/10.1002/sim.7208

57. Royston P, Parmar MKB. The use of restricted mean survival time to estimate the treatment effect in randomized clinical trials when the proportional hazards assumption is in doubt. Stat Med [Internet]. 2011 Aug 30;30(19):2409–21. Available from: https://onlinelibrary.wiley.com/doi/10.1002/sim.4274

58. Royston P, Parmar MK. Restricted mean survival time: an alternative to the hazard ratio for the design and analysis of randomized trials with a time-to-event outcome. BMC Med Res Methodol [Internet]. 2013 Dec 7;13(1):152. Available from: https://bmcmedresmethodol.biomedcentral.com/articles/10.1186/1471-2288-13-152

59. Mozumder SI, Rutherford MJ, Lambert PC. Estimating restricted mean survival time and expected life-years lost in the presence of competing risks within flexible parametric survival models. BMC Med Res Methodol [Internet]. 2021 Dec 11;21(1):52. Available from: https://bmcmedresmethodol.biomedcentral.com/articles/10.1186/s12874-021-01213-0

60. White IR, Royston P, Wood AM,. Multiple imputation using chained equations: issues and guidance for practice.. 4th ed. Vol. 30. 2011. 377–399 p.

61. World Medical Association Declaration of Helsinki. JAMA. 2013 Nov 27;310(20):2191.

62. Lubbe AL, Van Rijn M, Groen WG, Hilhorst S, Burchell GL, Hertogh CMPM, et al. The quality of geriatric rehabilitation from the patients’ perspective: a scoping review. Age Ageing. 2023 Mar 1;52(3).

63. van der Wouden JC, Blankenstein AH, Huibers MJH, van der Windt DAWM, Stalman WAB, Verhagen AP. Survey among 78 studies showed that Lasagna’s law holds in Dutch primary care research. J Clin Epidemiol [Internet]. 2007 Aug;60(8):819–24. Available from: https://linkinghub.elsevier.com/retrieve/pii/S0895435606004343

